# Optimising bi-level non-invasive ventilation in preterm neonates: a systematic review

**DOI:** 10.1101/2023.02.22.23286297

**Authors:** Jack Pickard

## Abstract

Bi-level non-invasive ventilation (BiPAP) can be used as a step-up from continuous positive airway pressure (CPAP) in preterm neonates to reduce the amount of time spent mechanically ventilated. Prolonged mechanical ventilation is associated with increased morbidity and mortality. MEDLINE was searched using the terms CPAP and BiPAP. Four studies reported a significant reduction in the need for mechanical ventilation when applying BiPAP compared with CPAP. Two studies reported no significant benefit. Studies which used 15/5 cm H_2_O or 20/5 cm H_2_O were more successful than those that used 6/5 cm H_2_O or 8/5 cm H_2_O. There was no discernible pattern to the effectiveness of respiratory rate, synchronisation or inspiratory time. In conclusion, BiPAP should be delivered at 15-20/5 cm H_2_O or 20/5 cm H_2_O.

**Key messages:** BiPAP has greater efficacy than CPAP at reducing the need for mechanical ventilation in preterm neonates with respiratory distress

An inspiratory pressure of at least 15 cm H_2_O should be employed wherever possible

There is insufficient evidence to recommend any particular respiratory rate, inspiratory time or synchronisation mode over another

**Structured clinical question:** Is BiPAP (intervention) more effective than CPAP (control) at reducing the need for mechanical ventilation in preterm neonates, and if so, what are the most effective pressures, inspiratory time, respiratory rate and synchronization mode to use?

**Search strategy:** MEDLINE was searched via Pubmed using the terms ‘CPAP’ AND ‘BiPAP’. This yielded 223 results. Further references within these articles were considered. Studies were included if they compared the effect of BiPAP vs CPAP on the need for mechanical ventilation or tracheal intubation. A total of 18 relevant studies were identified, including 15 randomised controlled trials (RCT) and one meta-analysis. Eight studies were excluded because they were already reported in the meta-analysis. Two were excluded because they were retrospective. A further two were excluded due to a lack of statistical analysis in the reporting. [1, 2]. A total of six studies remained for consideration; see table.

## Summary

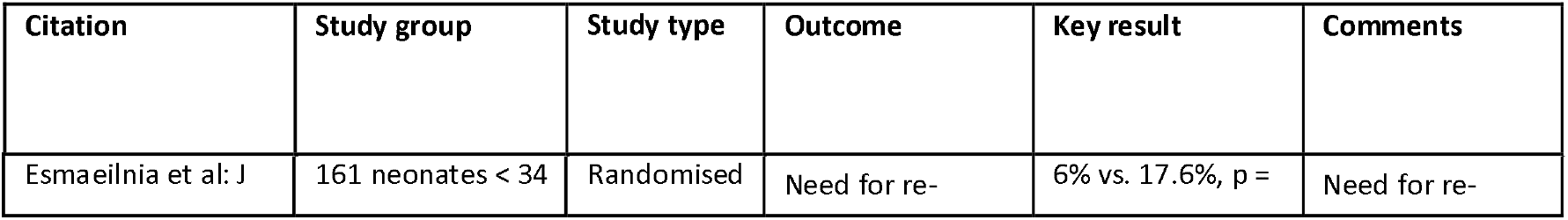
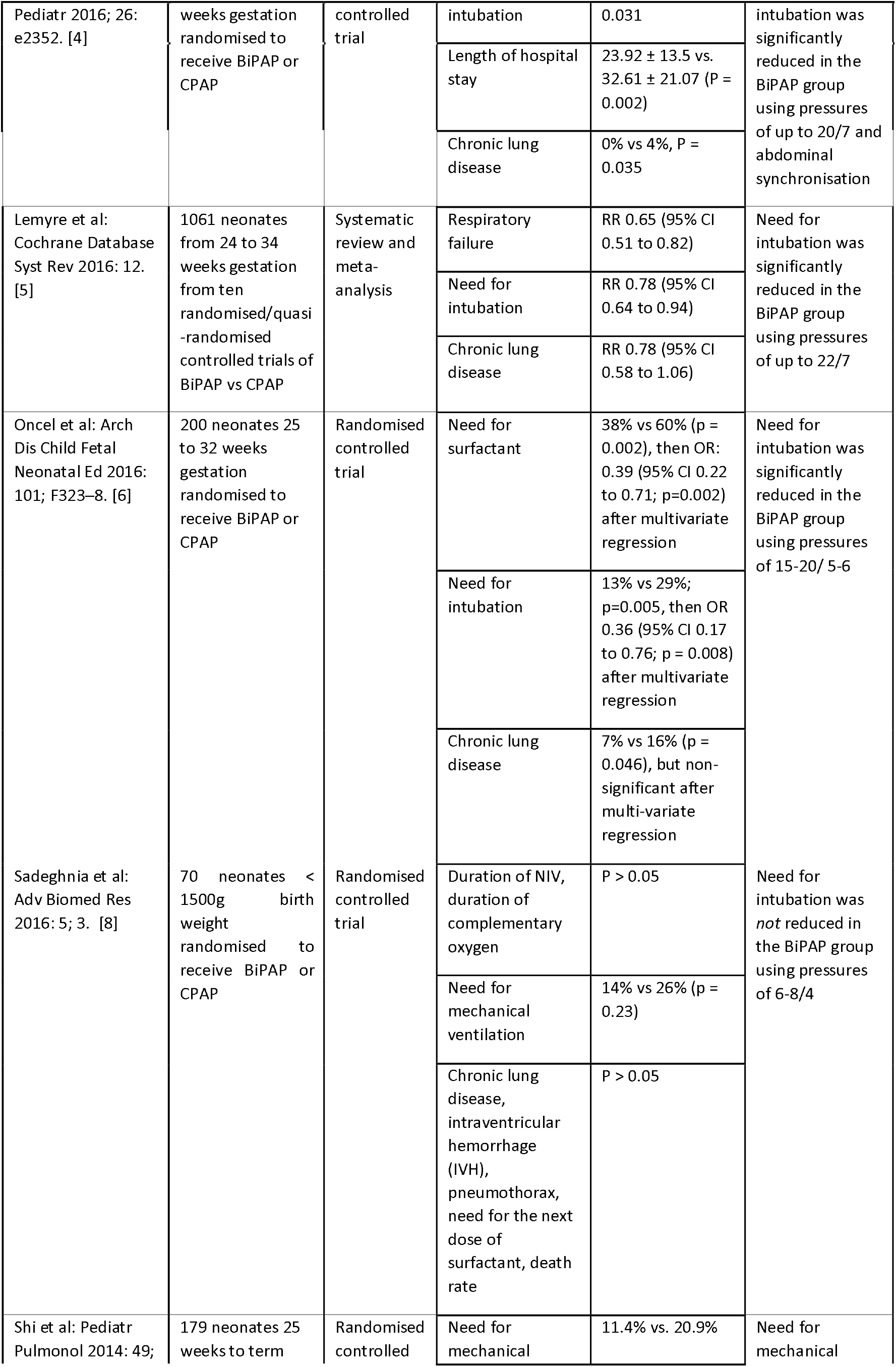
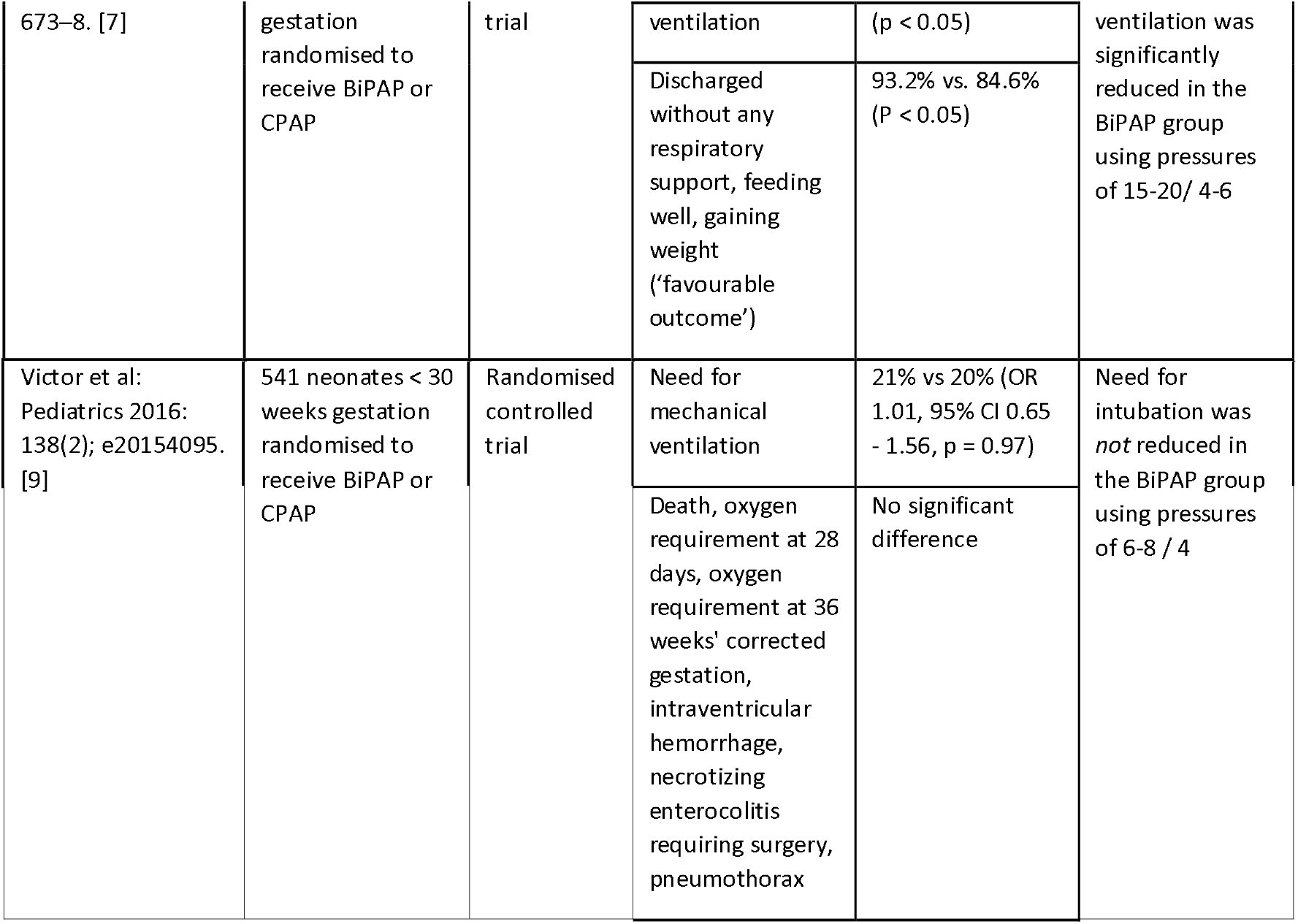

### Commentary

The primary purpose of non-invasive respiratory support (NIV) in preterm neonates, who frequently suffer with airway disease associated with early surfactant deficiency (respiratory distress syndrome; RDS), is to reduce the amount of time that is spent mechanically ventilated. Prolonged mechanical ventilation is associated with an increased risk of bronchopulmonary dysplasia (BPD; also known as chronic lung disease or CLD) and a poorer neurological outcome. [3] The most commonly used modalities of NIV are continuous positive airway pressure (CPAP), biphasic positive airway pressure (BiPAP), and heated humidified high flow nasal cannula oxygen (HFNC). BiPAP, also known as Bi-Phasic, non-invasive positive pressure ventilation (NIPPV), non-invasive mechanical ventilation and non-invasive mandatory ventilation (NIMV), consists of the application of intermittently higher pressure over positive end-expiratory pressure (PEEP). BiPAP is typically used in preterm neonates who are deemed to have the highest risk of requiring tracheal intubation and mechanical ventilation. This represents an assumption that BiPAP has greater efficacy in clearing carbon dioxide, improving oxygenation, and/or reducing overall respiratory effort when compared to CPAP or HFNC. However, there is a lack of consensus regarding which pressures, trigger methods, inspiratory time and respiratory rate are most effective when BiPAP is delivered, and indeed whether BiPAP truly does confer enhanced respiratory benefit. HFNC is used less commonly in extreme preterm neonates who are most likely to require mechanical ventilation and will not be considered in this review.

Four studies, including the Cochrane collaboration meta-analysis, reported a significant reduction in the need for mechanical ventilation when applying BiPAP compared with CPAP. [4, 5, 6, 7] Two studies reported no significant benefit. [8, 9] The largest, highest quality study (the Cochrane meta-analysis) found a relative risk of intubation of 0.78 (95% CI 0.64 to 0.94). [5] Within the Cochrane review, the single study which individually posted a significant benefit, Sai Sai Sunil Kishore et al [10], used pressures of 15-16/5 cm H_2_O. The most striking difference about the two studies that did not report a benefit, Sadeghnia [8] and Victor, [9] was that they used much lower pressures than the other studies (6-8/4 cm H_2_O). It is notable that CPAP was used at between 4 and 8 cm H_2_O in all studies, typically achieving a lower mean airway pressure (MAP) than that of BiPAP. There is an argument that the comparison is therefore not a fair one; future research should investigate the application of CPAP with a higher MAP.

One study used synchronisation of pressure cycling with the respiratory pattern of the neonate, timed with an abdominal sensor. [4] They achieved a positive result, however the remainder of studies used an unsynchronized, variable or unreported format and achieved mixed results. A wide variety of respiratory rates, from 10 to 50 breaths per minute, were employed across studies with no discernible relationship to the outcomes that were achieved. Inspiratory time was only reported by Sadeghnia et al, [8] who used 0.5 seconds.

The development of BPD / CLD was examined in several studies. While Esmaeilnia and Sadeghnia [4, 8] found significant reductions, this was not present in the remainder of studies. The Cochrane meta-analysis showed only a trend towards a benefit (RR 0.78, 95% CI 0.58 to 1.06). [5] BiPAP was regarded as safe in all studies, even when high inspiratory pressures of as much as 22 cm H_2_O were employed. One study reported traumatization of nasal skin and mucosa being a problem in both groups, [4] but this recovered fully in all cases.

In conclusion, BiPAP has greater efficacy than CPAP for reducing the need for mechanical ventilation in preterm neonates. Since higher inspiratory pressures tend to be more effective and their safety has been widely demonstrated, BiPAP should be applied at higher pressures wherever possible, with the upper pressure set at a minimum of 15 cm H_2_O. Pressures of up to 22 cm H_2_O are acceptable. There is insufficient evidence at present to say which respiratory rate, inspiratory time or synchronization mode is most effective for the delivery of BiPAP in preterm neonates.

## Data Availability

Review article so not relevant

## Notes

### Competing Interest Statement

The authors have declared no competing interest.

### Funding Statement

None

